# Comprehensive analysis of key m6A RNA modification-related genes and immune infiltrates in hypertrophic cardiomyopathy

**DOI:** 10.1101/2024.11.14.24317129

**Authors:** Xia Hu, Bo Liang

## Abstract

Hypertrophic cardiomyopathy (HCM) is the most common inherited heart disease. We performed a comprehensive analysis to construct the correlation of m6A and immune in HCM. Two HCM datasets (GSE141910 and GSE160997) and m6A-related regulators were obtained from GEO and published articles, respectively. Differentially expressed m6A-related regulators were obtained. Random forest model and nomogram were conducted to assess the risk of HCM, and finally, the m6A subtype was constructed. Functional enrichment analysis was conducted. Protein-protein interaction network of differentially expressed genes between m6A subtypes was performed. Furthermore, we constructed the Hubgene-chemical network, Hubgene-microRNA network, and Hubgene-transcription factor network of the top 10 hubgenes. Additionally, the immune subtype and hubgene subtype were constructed. PCR was performed to validate the m6A-related regulators. We obtained 20 m6A-related regulators in HCM. Among them, 8 m6A-related regulators differentially expressed (YTHDC1, HNRNPC, and FMR1 were up-regulated while YTHDC2, FTO, WTAP, IGF2BP2, and IGF2BP3 were down-regulated). FTO, FMR1, IGF2BP3, YTHDC1, and IGF2BP2 were the top 5 important m6A-related regulators and were used to conduct the nomogram. We obtained 329 differentially expressed genes in m6A subtype and these genes enriched HCM-related processes and pathways. Furthermore, we constructed the Hubgene-chemical network, Hubgene-microRNA network, and Hubgene-transcription factor network of the top 10 hubgenes (NFKBIA, NFKB1, PSMA3, PSMC4, PSMA2, PSMA4, PSMD7, PSMD10, PSMD8, and PSMA6). And then we constructed an immune subtype based on the immune cell infiltration levels and hubgene subtype based on the expression of the top 10 hubgenes. Finally, we verified the main results through experiments. In conclusion, we built a nomogram and identified 8 m6A-related regulators and 10 hubgenes, which were prominently associated with HCM. We found that m6A and the immune system may play a crucial role in the HCM. Accordingly, those genes and pathways might become therapeutic targets with clinical usefulness in the future.

## 1. Introduction

Hypertrophic cardiomyopathy (HCM) is the commonest primitive inherited disease of the myocardium[1], with an autosomal dominant pattern of inheritance and a worldwide prevalence of approximately one in 500 adult subjects[2, 3]. HCM is defined principally by left ventricular hypertrophy without increased cardiac afterload or another underlying pathophysiological etiology, and causes heart failure at any age[4]. A clinical diagnosis of HCM in adults can be established by imaging with 2D echocardiography or cardiovascular magnetic resonance[5]. According to the latest guidelines[2, 3], pharmacological therapy for HCM patients includes non-selective drugs such as β-blockers, cardiac-selective calcium antagonists, and disopyramide, to be used in symptomatic patients with obstruction of the left ventricular outflow tract. Current drugs are unable to address the pathophysiological mechanisms of left ventricular dysfunction in HCM and are not, therefore, effective in preventing arrhythmias or slowing down disease progression in HCM patients[1]. Novel allosteric inhibitors of myosin are being developed and clinically validated, specifically targeting HCM-related pathophysiological mechanisms, such as myocardial hypercontractility and altered energetics[6]. However, HCM is a heterogeneous disease that involves numerous sarcomere-independent morphological features, suggesting that the precise mechanisms of HCM have not been fully elucidated.

Epigenetic dysregulation is crucial for the pathological development of cardiovascular diseases[7, 8]. N6-methyladenosine (m6A) is the most common post-transcriptional modification of mRNA[9] and has recently attracted great attention. m6A influences a series of biological functions in cardiovascular diseases. m6A RNA modification mediates the atherogenic inflammatory cascades in vascular endothelium[10] and modulates endothelial atherogenic responses to disturbed flow in mice[11]. Age-related differences in m6A in response to acute myocardial ischemia/reperfusion injury[12] and m6A might play important roles in blood pressure regulation[13]. m6A modification effectively improves the cardiac function and decreases the infarct size in acute myocardial infarction through tricarboxylic acid cycle-based metabolic reprogramming[14] and promotes miR-133a repression during cardiac development and hypertrophy via IGF2BP2[15]. In heart failure, FTO-dependent cardiac m6A methylome plays a functional importance in cardiac contraction[16] and m6A might be an interesting target for therapeutic interventions[17]. However, the role of m6A in HCM is largely unknown.

m6A RNA modification acts as a novel regulator in both innate and adaptive immune response[18, 19]. In acute myocardial infarction and aortic dissection, m6A is closely connected to immune cell infiltration[20, 21]. Previous study showed that the changes in the immune system engage in the HCM mechanisms[22, 23] and the systemic immune-inflammation index is a significant risk factor for all-cause mortality in HCM patients[24]. All these evidence supports immune response has a significant impact on the HCM biology and outcome. However, the crosstalk between m6A and immune response in HCM is poorly understood. In this study, we comprehensively evaluated the expression of m6A-related regulators and immune cell infiltration in HCM patients from two independent cohorts, constructed random forest model and nomogram, and further identified m6A and immune subtype (Figure 1).

**Figure 1.**
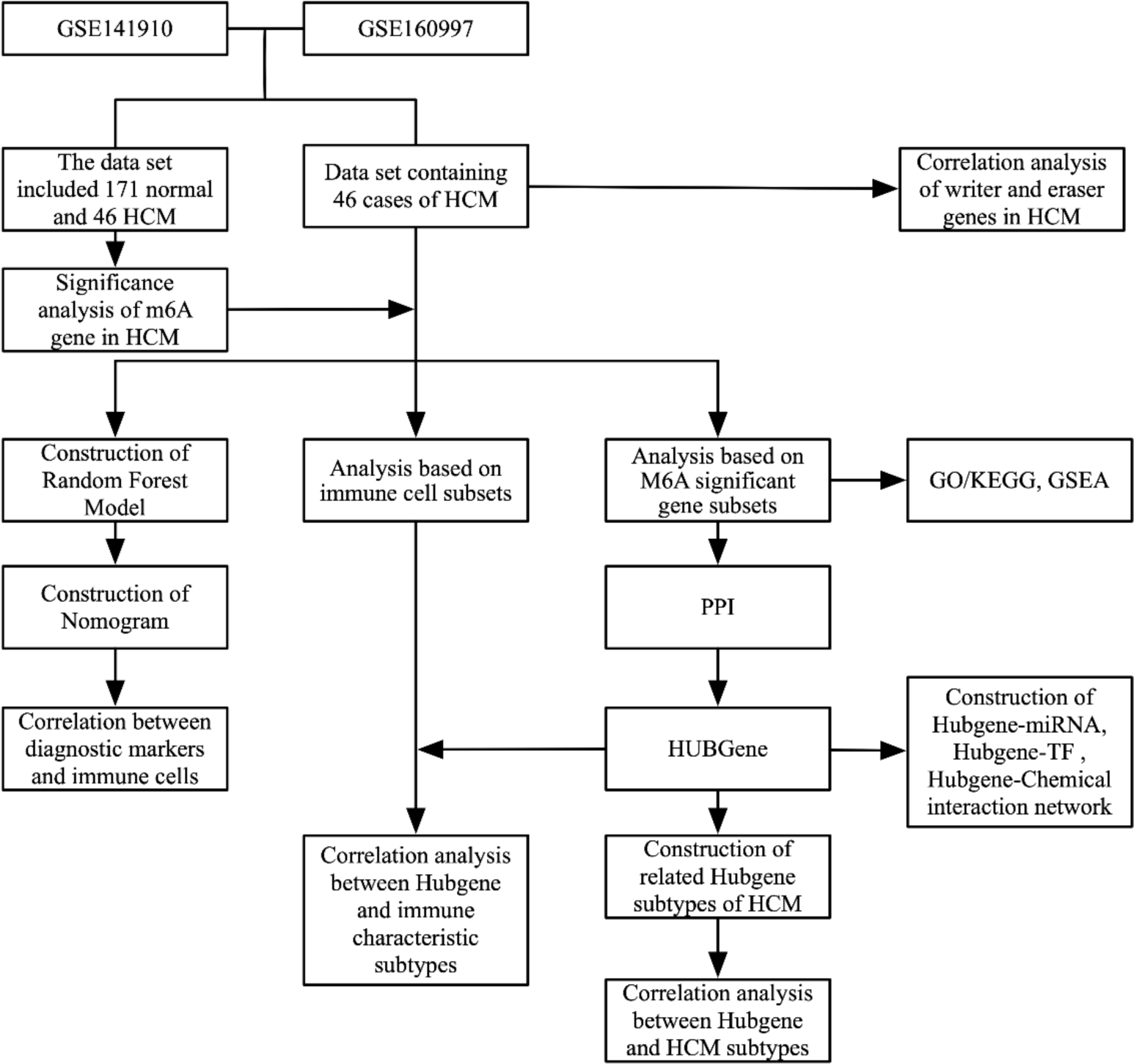
Flow chart of this study.

## 2. Methods

### 2.1 Data collection

Two HCM datasets from GEO, namely GSE141910 and GSE160997[25], were obtained though the *GEOquery* package and details are shown in Table 1. All samples were pooled after removing batch effects (Figure S1), as descried previously[26, 27]. In addition, m6A-related regulators were extracted based on published literature.

### 2.2 m6A-related regulators in HCM

The chromosomal locations of m6A-related regulators were visualized using the *RCircos* package and the differential expression levels of m6A-related regulators between HCM patients and the controls were analyzed using the *limma* package. Finally, linear regression analysis was used to explore the correlation between m6A writers and m6A erasers.

### 2.3 Construction of random forest model and nomogram

Random forest is a compositionally supervised learning method that can be viewed as an extension of decision tree[28]. We divided patients with HCM into the training cohort and the validation cohort by 7:3, and selected candidate m6A regulators from all m6A-related regulators to predict the occurrence of HCM. We built the random forest model in the training cohort with ntrees = 500, and then analyzed the importance of m6A-related regulators and evaluated the model with residual analysis and ROC curves on both training and validation cohorts.

We constructed a nomogram based on the top 5 candidate m6A regulators using the *rms* package to predict the prevalence of HCM patients. Calibration curve, decision curve analysis, and ROC curve were conducted to evaluate the nomogram, as described previously[29, 30]. Finally, we explored the correlation between top 5 candidate m6A regulators and immune-related cell types.

### 2.4 m6A subtype construction

We used the *ConsensusClusterPlus* package[31] to identify the membership and number of clusters in datasets based on the expression of the top 5 m6A-related regulators. Then individuals were clustered into corresponding clusters. We further analyzed the expression of the top 10 hubgenes in corresponding clusters. The differentially expressed genes among clusters were analyzed using the *limma* package with a threshold of adjusted *P* < 0.05 and |log_2_Fold Change| > 2 and the *ggplot2* package[26] was used to visualization.

We used the *clusterProfiler* package[32] to perform gene ontology (including cellular component, molecular function, and biological process) and Kyoto Encyclopedia of Genes and Genomes enrichment analysis on differentially expressed genes with a threshold of *P* < 0.5 and false discovery rate < 0.05. The specific Kyoto Encyclopedia of Genes and Genomes terms were visualized through the *Pathview* package[33]. Gene sets of c2.cp.v7.4.symbols.gmt, which are curated from various sources, including online pathway databases and the biomedical literature, were downloaded from Molecular Signatures Database[34] as background for gene set enrichment analysis[35]. Adjusted *P* < 0.05 was considered statistically significant.

The STRING database[36] was used for protein-protein interaction analysis of differentially expressed genes, and those genes with a score greater than 0.6 were selected for later analysis. The *cytoHubba* plugin[37] in Cytoscape[38] was used to functionally enrich the cluster from the protein-protein interaction network to obtain the hubgenes. Furthermore, we constructed Hubgene-chemical network, Hubgene-microRNA network, and Hubgene-transcription factor network of the top 10 hubgenes obtained based on the protein-protein interaction network. NetworkAnalyst[39] was used to build Hubgene-chemical network and Hubgene-chemical network, and miRTarBase[40] was used to build Hubgene-microRNA network. Cytoscape[38] was used for visualization.

### 2.5 Immune subtype construction

We calculated the immune cell infiltration levels using the *ssGSEA* function of the *GSVA* package to obtain the enrichment score for each immune-related cell type. Subtype classification was further constructed based on the enrichment score features of immune-related cell types using the *ConsensusClusterPlus* package[31]. Then individuals were clustered into corresponding clusters. The differentially expressed genes among clusters were analyzed using the *limma* package with a threshold of adjusted *P* < 0.05 and |log_2_Fold Change| > 2 and the *ggplot2* package[26] was used to visualization. We further analyzed the difference and correlation between top 10 hubgenes and immune infiltration.

CIBERSORT is a method for characterizing cell composition of complex tissues from their gene expression profiles[41]. We obtained immune cell infiltration matrix by CIBERSORT with *P* < 0.05 as threshold. The *ggplot2* package[26] and *corrplot* package were used to visualization. We further analyzed the correlation between top 10 hubgenes and immune cells.

### 2.6 Hubgene subtype construction

Subtype classification was constructed based on the expression of top 10 hubgenes using the *ConsensusClusterPlus* package[31]. Then individuals were clustered into corresponding clusters. We further analyzed the expression of top 10 hubgenes in corresponding clusters.

### 2.7 Experimental Validation

The 8-week-old C57BL/6J mice were obtained from Vital River (Beijing, China) and all animal experiments were reviewed and approved by the Experimental Animal Center of Zhengzhou University (Zhengzhou, China) (ZZU-LAC20220311[23]). We conducted transverse aortic constriction (TAC) to build the myocardial hypertrophy model[42]. A total of 12 male mice were divided into the Sham group (N = 6) and the TAC group (N = 6). After four weeks of TAC, the mice were sacrificed after transthoracic echocardiography for the following experiments. Histomorphological changes in the heart were determined by HE and WGA staining according to our previous protocol[43, 44]. The mRNA expression of atrial natriuretic peptide (Anp), brain natriuretic peptide (Bnp), myosin heavy chain 7 (Myh7), Hnrnpc, Fmr1, and Fto in the heart tissue was quantified by real-time quantitative polymerase chain reaction assay according to our previous protocol[43, 44]. The primer sequences are listed in Table S1.

### 2.8 Statistical analysis

The experimental data are expressed as the mean and standard deviation and were analyzed by Prism (version 9.5.0, GraphPad, CA, USA) with a t test and one-way analysis of variance if applicable. P < 0.05 was considered statistically significant.

All data processing and analysis were completed through R (version 4.1.0) with independent Student t test, Mann Whitney U test (Wilcoxon rank sum test), Chi square test or Fisher exact test, if applicable. All *P* < 0.05 bilaterally was considered statistically significant.

## 3. Results

### 3.1 m6A-related regulators in HCM

We obtained 20 m6A-related regulators in HCM. Among them, there were 6 writers (METTL3, ZC3H13, METTL14, CBLL1, WTAP, and RBM15), 2 erasers (FTO and ALKBH5), and 12 readers (YTHDC1, YTHDC2, ELAVL1, YTHDF1, LRPPRC, YTHDF2, FMR1, YTHDF3, HNRNPC, HNRNPA2B1, IGF2BP2, and IGF2BP3). The chromosomal locations of 20 m6A-related regulators were shown in Figure 2A. A total of 8 m6A-related regulators differentially expressed, YTHDC1, HNRNPC, and FMR1 were up-regulated in HCM and YTHDC2, FTO, WTAP, IGF2BP2, and IGF2BP3 were down-regulated in HCM (Figure 2B&C). Through linear regression analysis, we found that the expression of CBLL1, ZC3H13, METTL3, and METTL14 was highly positively correlated with FTO (*P* < 0.05, < 0.0001, < 0.0001, and < 0.0001, respectively), the expression of RBM15 was highly negatively correlated with FTO (*P* < 0.0001), and the expression of CBLL1 and METTL14 was highly negatively correlated with ALKBH5 (*P* < 0.001 and < 0.01, respectively) (Figure 2D).

**Figure 2.**
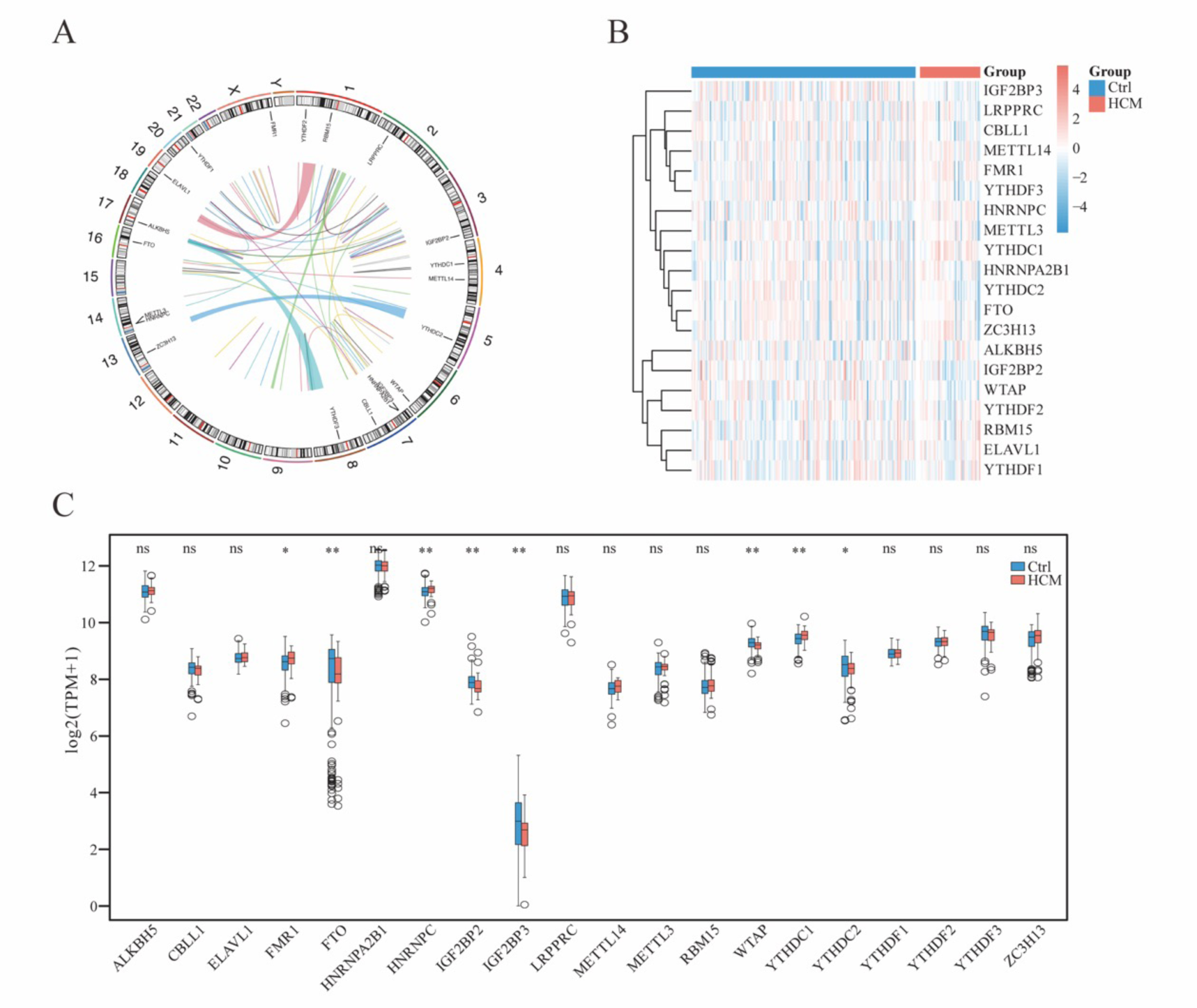
The landscape of m6A-related regulators in HCM. A. The chromosomal locations of 20 m6A-related regulators. B. Heatmap of expression of 20 m6A-related regulators. C. Histogram of expression of 20 m6A-related regulators. \**P* < 0.05, \*\**P* < 0.01, \*\*\**P* < 0.001. D. The correlation between writers and erasers.

### 3.2 Random forest model and nomogram

In the training cohort, we build the random forest model (Figure 3A). FTO, FMR1, IGF2BP3, YTHDC1, and IGF2BP2 were top 5 important m6A-related regulators in HCM (Figure 3B). The residual plots in the training and validation cohorts were shown in Figure 3C&D (*P* < 0.0001 and < 0.001, respectively). Moreover, The ROC curves indicated that the model performed well (AUC = 1 and 0.86 in the training and validation cohorts, respectively) (Figure 3E).

**Figure 3.**
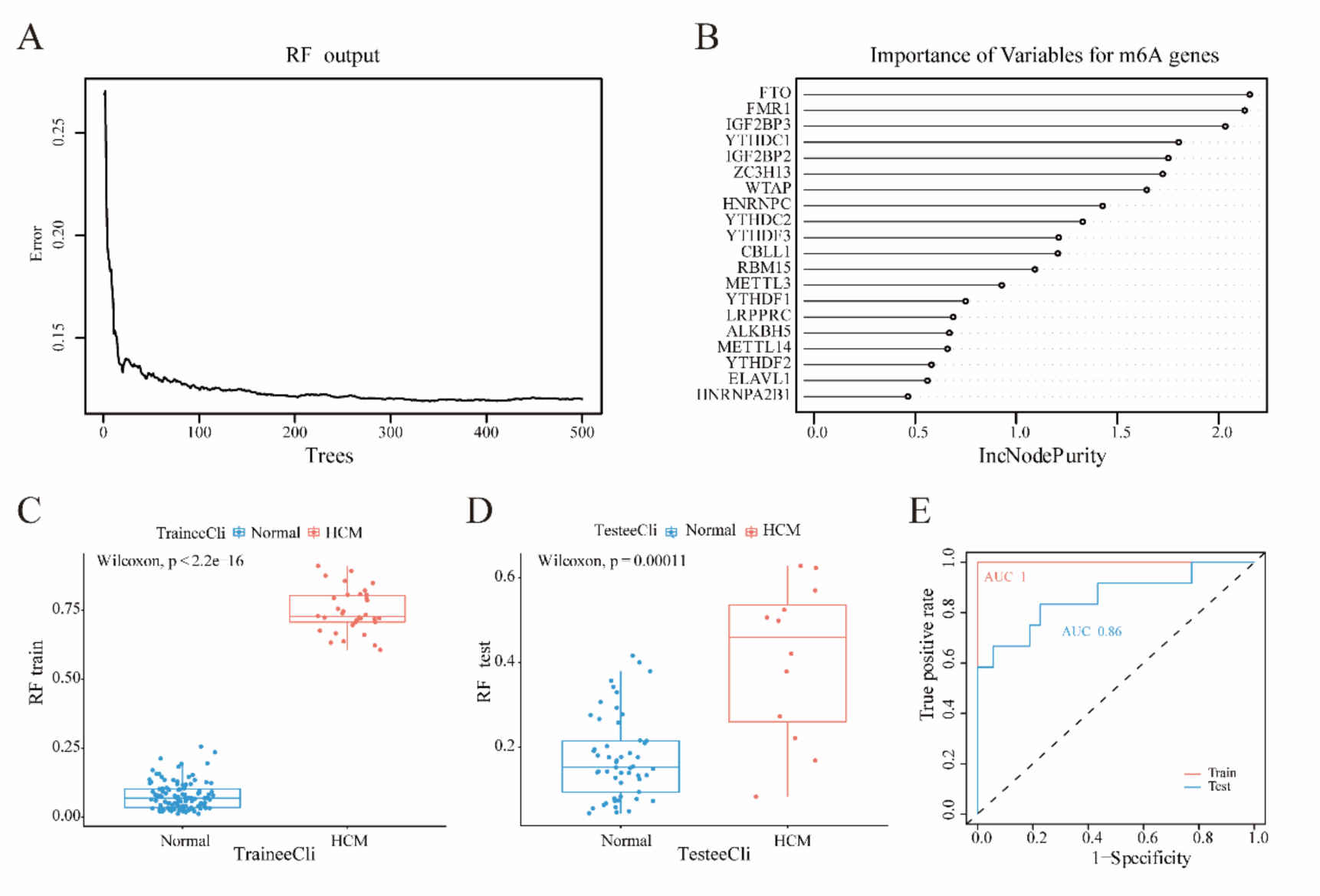
Random forest model construction. A. Model was built in the training cohort. B. Importance of m6A-related regulators based on the model. C. Residual plot showing the distribution of residuals in the training cohort. D. Residual plot showing the distribution of residuals in the validation cohort. E. The ROC curves showing the accuracy of the model in the training and validation cohorts.

We constructed a nomogram based on top 5 important m6A-related regulators in HCM (Figure 4A) and calibration curve, decision curve analysis, and ROC curve showed that the performance of nomogram was acceptable (Figure 4B∼D).

**Figure 4.**
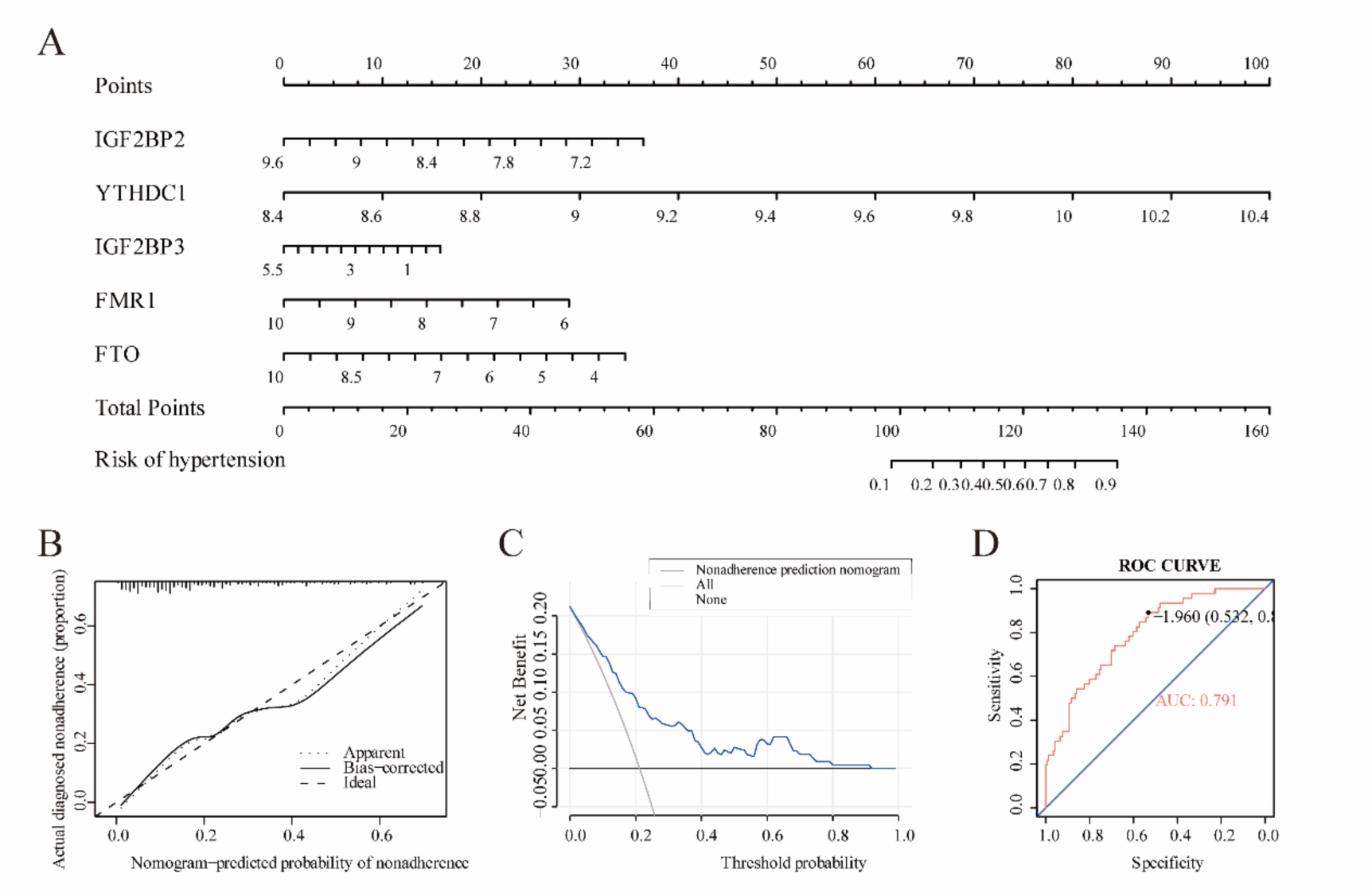
Construction of the nomogram. A. Construction of the nomogram based on top 5 important m6A-related regulators in HCM. B. Calibration curve. C. decision curve analysis. D. ROC curve.

Correlation analysis showed that FMR1 and T cells follicular helper, FTO and B cells naïve, Monocytes, T cells CD8, T cells regulatory (Tregs), IGF2BP2 and Dendritic cells resting, T cells gamma delta, T cells regulatory (Tregs), IGF2BP3 and Dendritic cells activated, Dendritic cells resting, T cells gamma delta, YTHDC1 were strongly correlated with T cells CD8 (Figure 4-1).

**Figure 4-1.** Correlation between top 5 candidate m6A regulators and immune-related 654 cell types.

### 3.3 m6A subtype construction

When k = 2, the classification was reliable and stable (Figure 5A∼F). Among 10 hubgenes, NFKB1, PSMC4, PSMA4, PSMA2, and PSMA6 were dys-regulated in ClusterA and ClusterB (*P* < 0.001, < 0.01, < 0.001, < 0.01, and < 0.001, respectively) (Figure 5). Therefore, we divided the samples into ClusterA and ClusterB. Further, we obtained 329 differentially expressed genes (154 genes were up-regulated and 175 genes were down-regulate) (Figure 5G&H).

**Figure 5.**
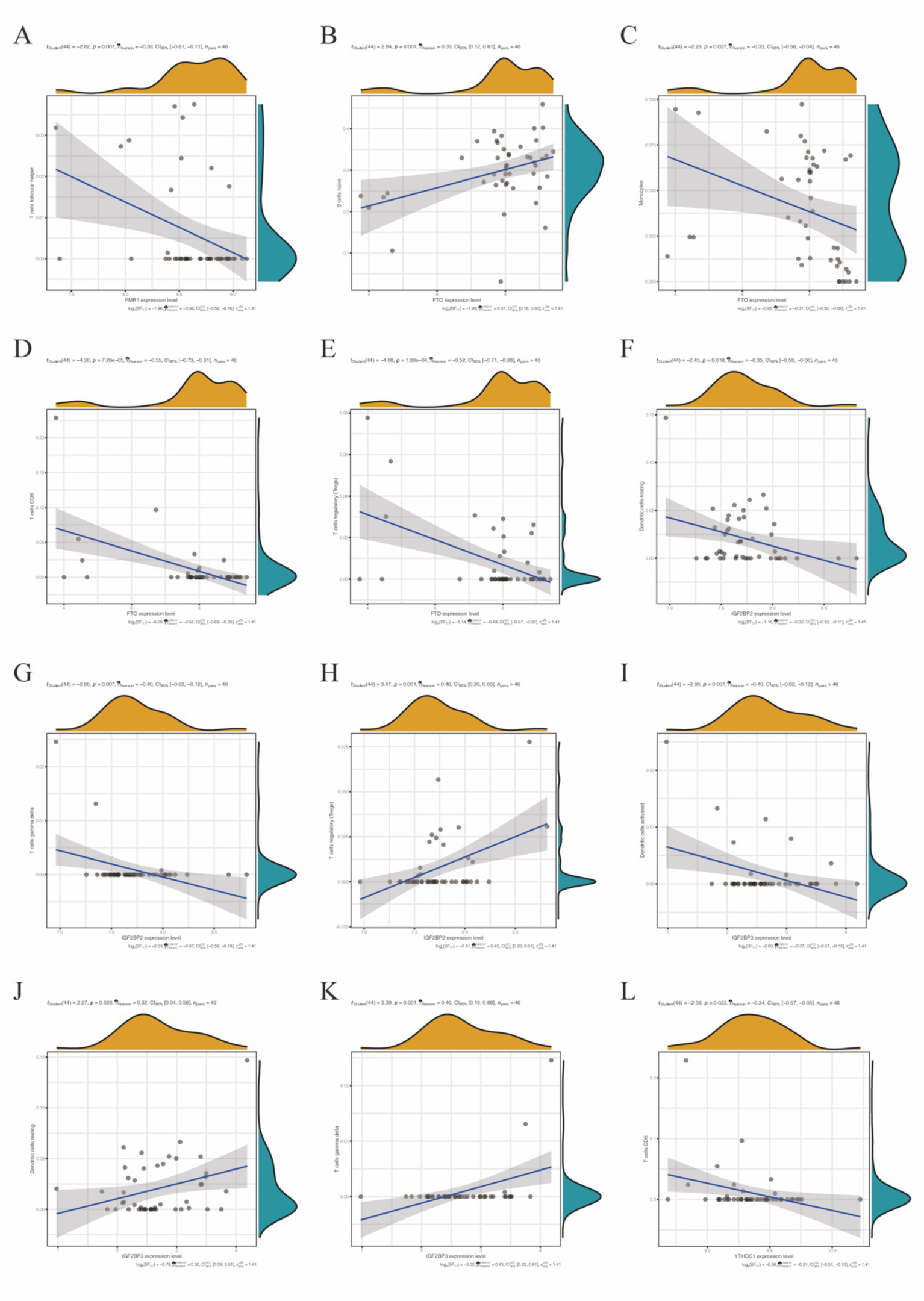
m6A subtype construction based on the expression of top 5 m6A-related regulators. A. Matrix heatmap for k = 2. B. Matrix heatmap for k = 3. C. Matrix heatmap for k = 4. D. Consistent cumulative distribution function plot. This figure allows a user to determine at what number of clusters, k, the cumulative distribution function reaches an approximate maximum, thus consensus and cluster con dence is at a maximum at this k. E. Delta area plot. The delta area score (y-axis) indicates the relative increase in cluster stability. F. Tracking plot. This plot provides a view of item cluster membership across different k and enables a user to track the history of clusters relative to earlier clusters. G. Heatmap for differentially expressed genes. H. Volcano plot for differentially expressed genes. I. Difference analysis of top 10 hubgene in m6A subtype classification.

Through enrichment analysis of 329 differentially expressed genes, we found that homophilic cell adhesion via plasma membrane adhesion molecules, positive regulation of necrotic cell death, respiratory electron transport chain, glycosylation, necrotic cell death, regulation of necrotic cell death, energy derivation by oxidation of organic compounds, mitochondrial ATP synthesis coupled electron transport, cellular response to osmotic stress, and positive regulation of amino acid transport were enriched in biological process; endocytic vesicle lumen, autolysosome, respiratory chain complex, Golgi cis cisterna, haptoglobin-hemoglobin complex, mitochondrial respirasome, mitochondrial respiratory chain complex IV, hemoglobin complex, Golgi cisterna membrane, and ficolin-1-rich granule membrane were enriched in cellular component; Rac GTPase binding, fibronectin binding, inorganic anion transmembrane transporter activity, intramolecular transferase activity, phosphotransferases, haptoglobin binding, RAGE receptor binding, pre-mRNA intronic binding, copper ion binding, oxygen binding, and oxygen carrier activity, were enriched in molecular function (Figure 6A). Calcium signaling pathway, Sphingolipid metabolism, Gastric acid secretion, Leukocyte transendothelial migration, Neuroactive ligand-receptor interaction, Malaria, and Proximal tubule bicarbonate reclamation were enriched in Kyoto Encyclopedia of Genes and Genomes (Figure 6B). Calcium signaling pathway and Leukocyte transendothelial migration were visualized in Figure 6C&D. Through gene set enrichment analysis, pathways related to infection, oxidative stress, immunity, and amino acid metabolism were enriched (Figure 6E).

**Figure 6.**
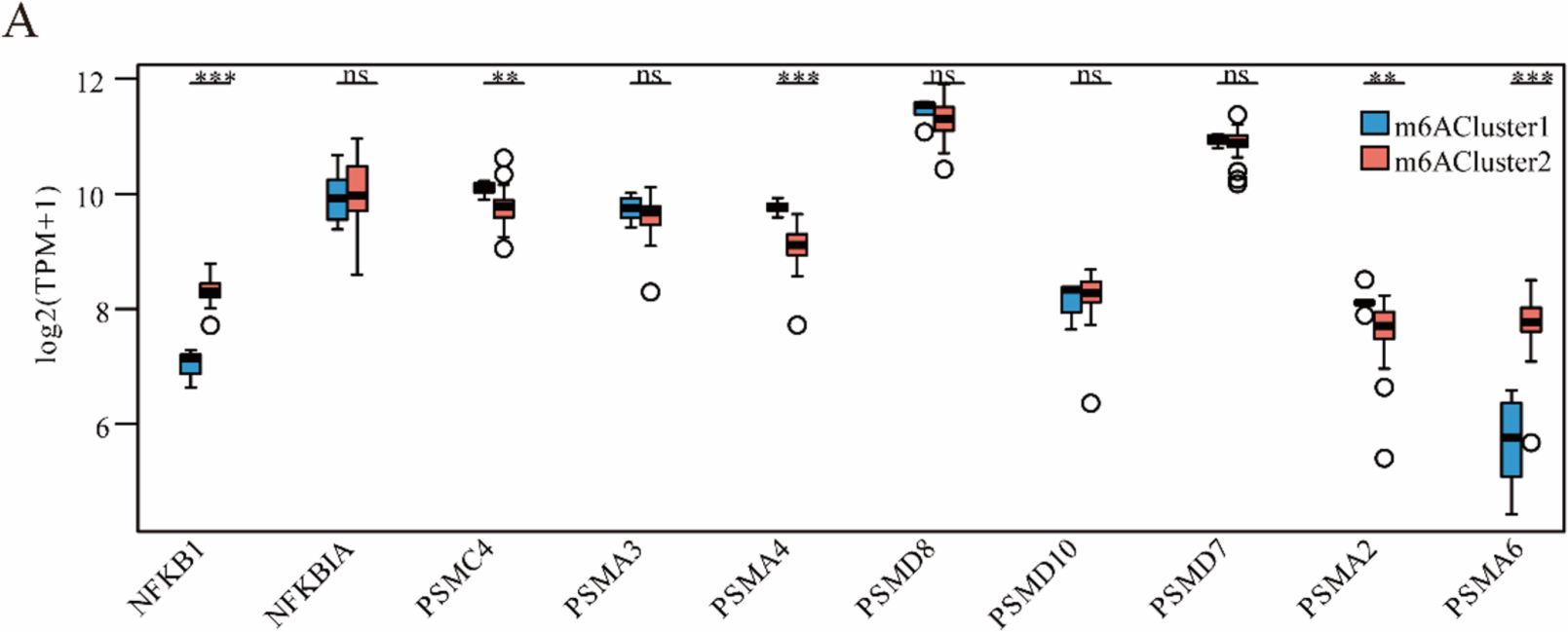
Functional enrichment of 329 differentially expressed genes. A. Top 10 gene ontology terms. B. Top 7 Kyoto Encyclopedia of Genes and Genomes terms. C. Visualization of Calcium signaling pathway. D. Visualization of Leukocyte transendothelial migration. E. Gene set enrichment analysis terms.

Through protein-protein interaction analysis of 329 differentially expressed genes, we obtained a network consisting of 143 genes and 184 connections (Figure 7A). Next, we got sub-networks consisting of the top 20 and 10 genes (Figure 7B&C).

**Figure 7.**
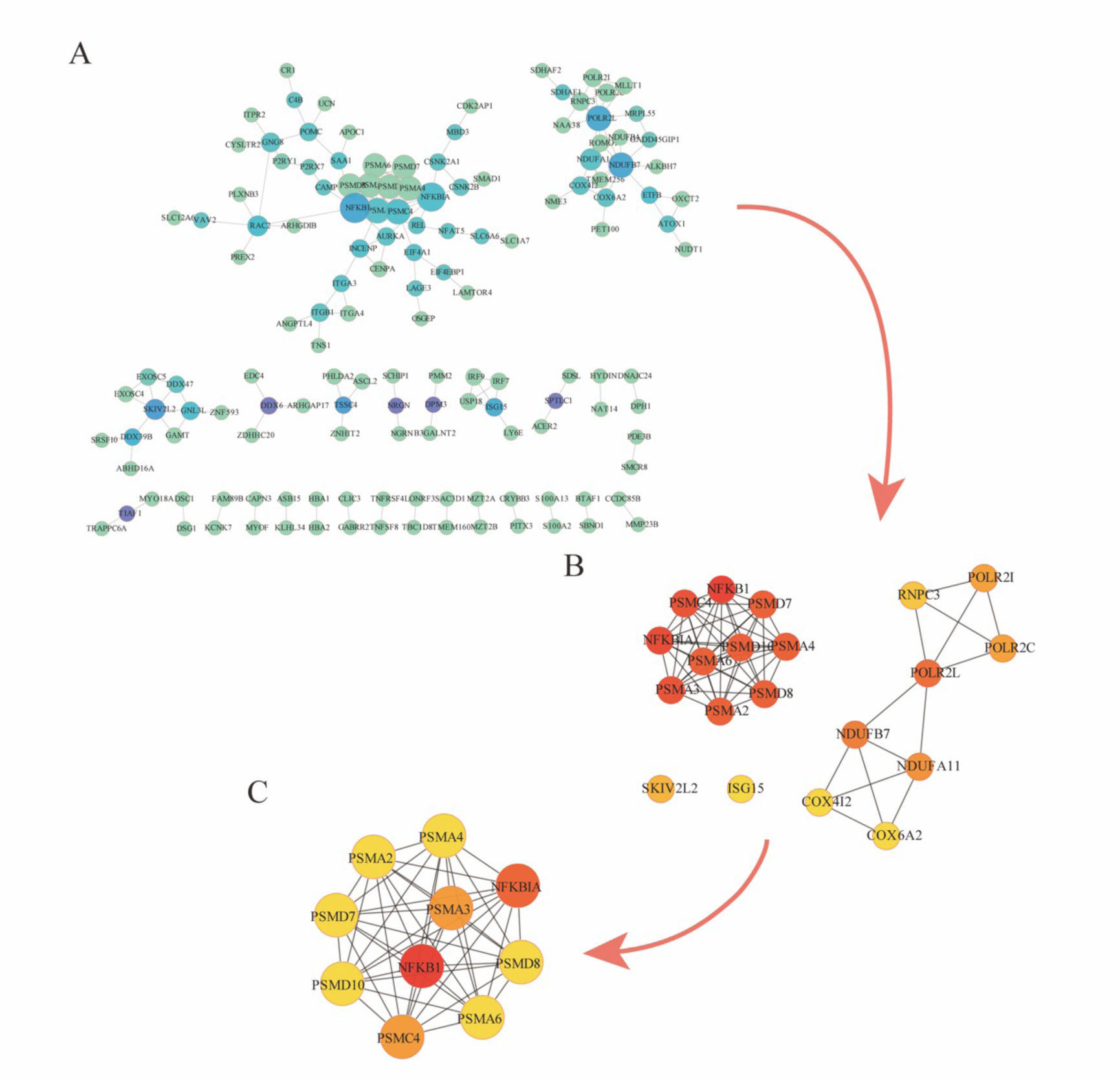
Protein-protein interaction network. A. Protein-protein interaction network among those genes with a score greater than 0.6. B. Protein-protein interaction network of top 20 hubgenes. C. Protein-protein interaction network of top 10 hubgenes.

Furthermore, we constructed Hubgene-chemical network, Hubgene-microRNA network, and Hubgene-transcription factor network of the top 10 hubgenes. Hubgene-chemical network included 574 nodes (10 hubgenes and 564 chemicals) and 837 edges (Figure 8A), Hubgene-microRNA network included 131 nodes (9 hubgenes and 121 microRNAs) and 132 edges (Figure 8B), Hubgene-transcription factor network included 181 nodes (10 hubgenes and 171 transcription factors) and 284 edges (Figure 8C).

**Figure 8.**
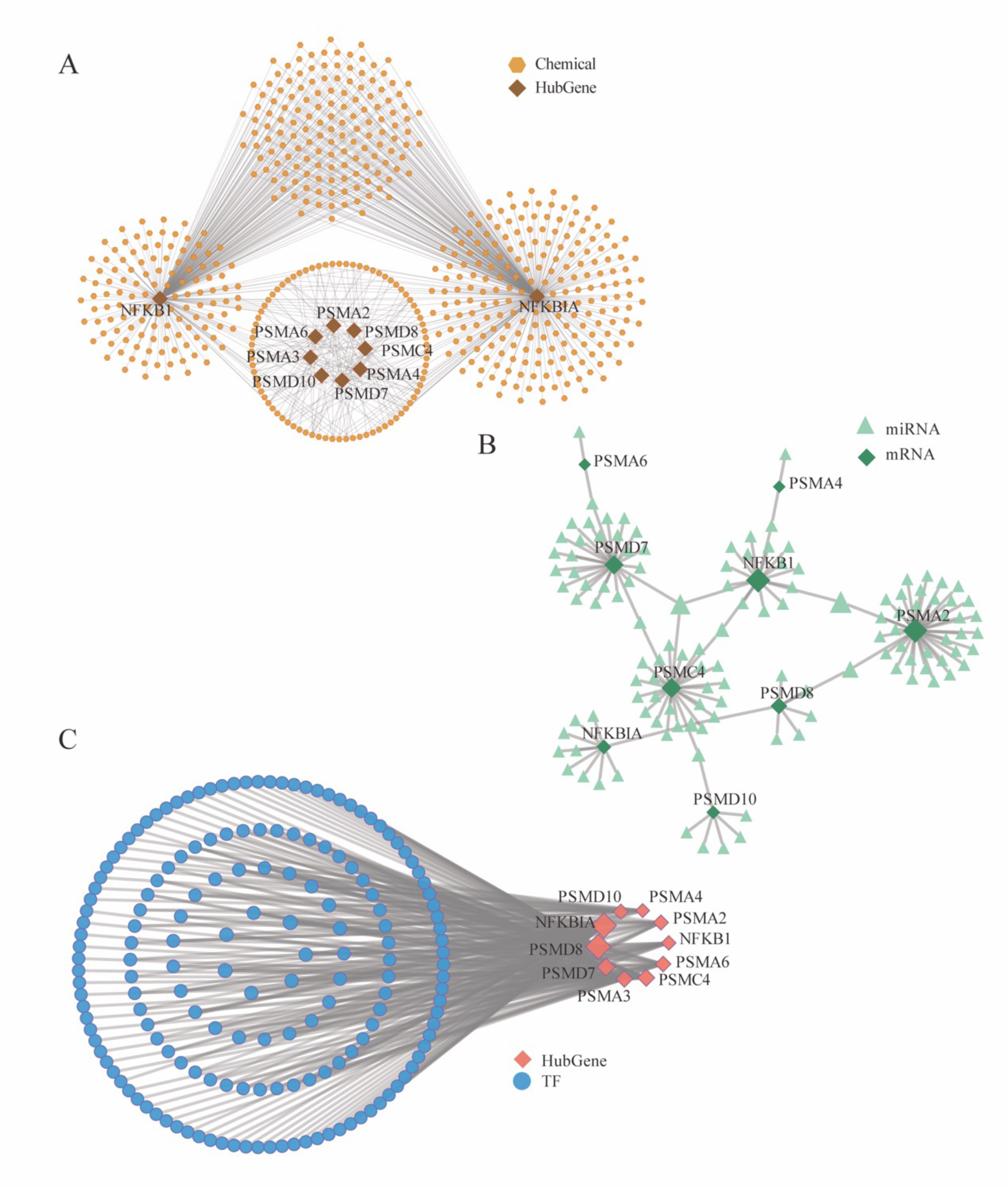
Networks of the top 10 hubgenes. A. Hubgene-chemical network. B. Hubgene-microRNA network. C. Hubgene-transcription factor network.

### 3.4 Immune subtype construction

When k = 2, the classification was reliable and stable (Figure 9A∼D). Therefore, we divided the samples into ImmuClusterA and ImmuClusterB. Further, we obtained 50 differentially expressed genes (15 genes were up-regulated and 35 genes were down-regulated) (Figure 9E). Among top 10 hubgenes, NFKB1, PSMD10, and PSMA6 negatively correlated with most immune cell infiltration and PSMC4 and PSMA4 positively correlated with most immune cell infiltration (Figure 9F). Interestingly, NFKB1, PSMD10, PSMA6, PSMC4, and PSMA4 were dys-regulated in ImmuClusterA and ImmuClusterB (*P* < 0.001, < 0.01, < 0.001, < 0.01, and < 0.01, respectively) (Figure 9G), suggesting these genes may be immune-related targets.

**Figure 9.**
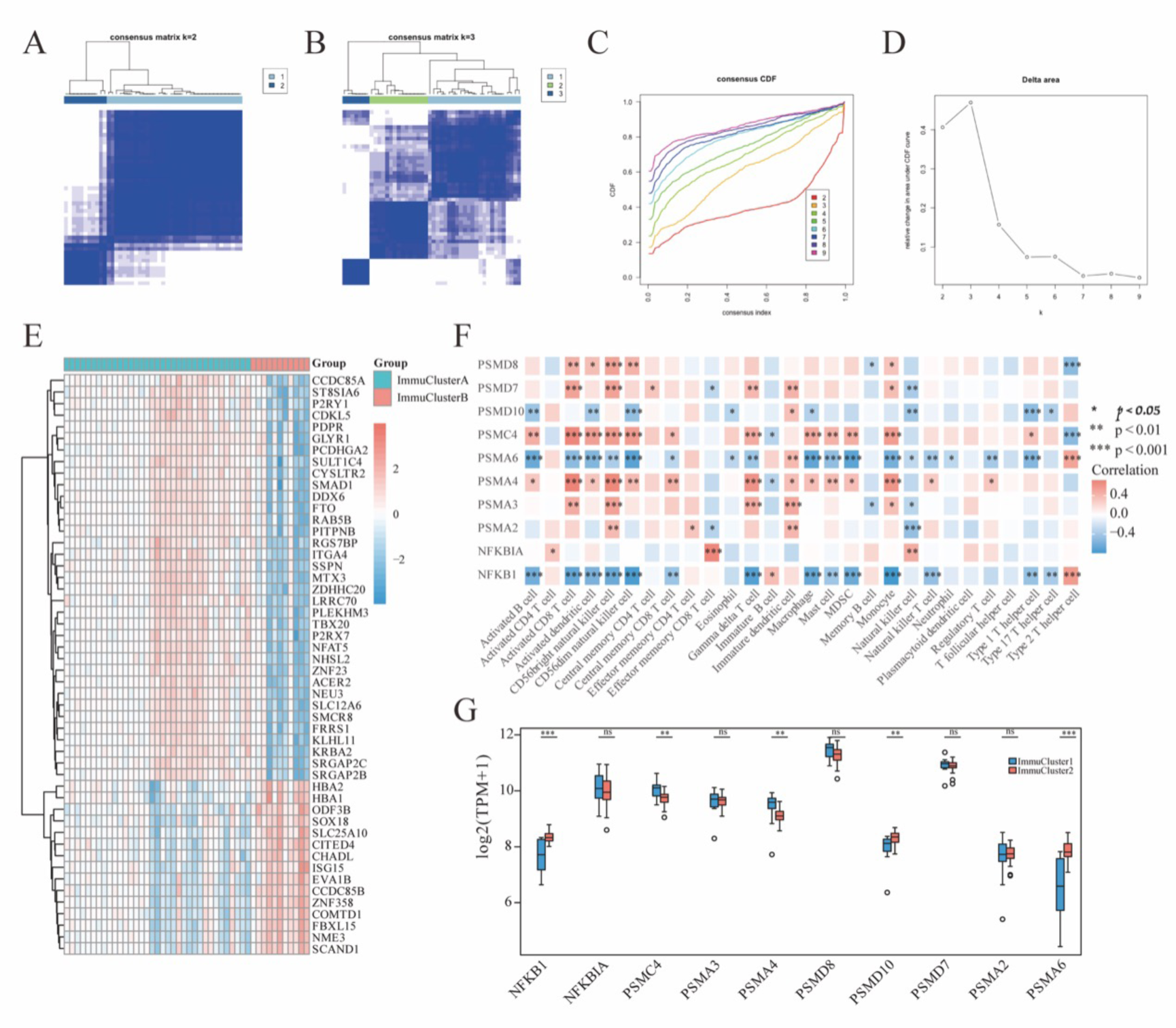
Immune subtype construction. A. Matrix heatmap for k = 2. B. Matrix heatmap for k = 3. C. Consistent cumulative distribution function plot. This figure allows a user to determine at what number of clusters, k, the cumulative distribution function reaches an approximate maximum, thus consensus and cluster con dence is at a maximum at this k. D. Delta area plot. The delta area score (y-axis) indicates the relative increase in cluster stability. E. Heatmap for differentially expressed genes. F. The correlation between top 10 hubgenes and immune-related cell types. G. Histogram of expression of top 10 hubgenes. \*\**P* < 0.01, \*\*\**P* < 0.001.

We used the CIBERSORT algorithm to analyze the proportion of immune subsets in patients with HCM, and constructed a map of 22 immune cells in HCM (Figure 10A) and the correlation of 22 immune cells (Figure 10B). We analyzed the correlation between top 10 hubgenes and immune cells, and found that NFKB1 was negatively correlated with CD8^+^ T cells and T regulatory cells (Figure 10 C&D), PSMA4 was negatively correlated with B naïve cells and positively correlated CD8^+^ T cells (Figure 10E&F), PSMA6 was negatively correlated with mast cells and T regulatory cells (Figure 10 G&H), PSMC4 was positively correlated T follicular helper cells and negatively correlated with T gamma delta cells (Figure 10 I&J), PSMD7 and PSMD8 were negatively correlated with T gamma delta cells (Figure 10 K&L) PSMD10 was negatively correlated with and T regulatory cells (Figure 10 M).

**Figure 10.**
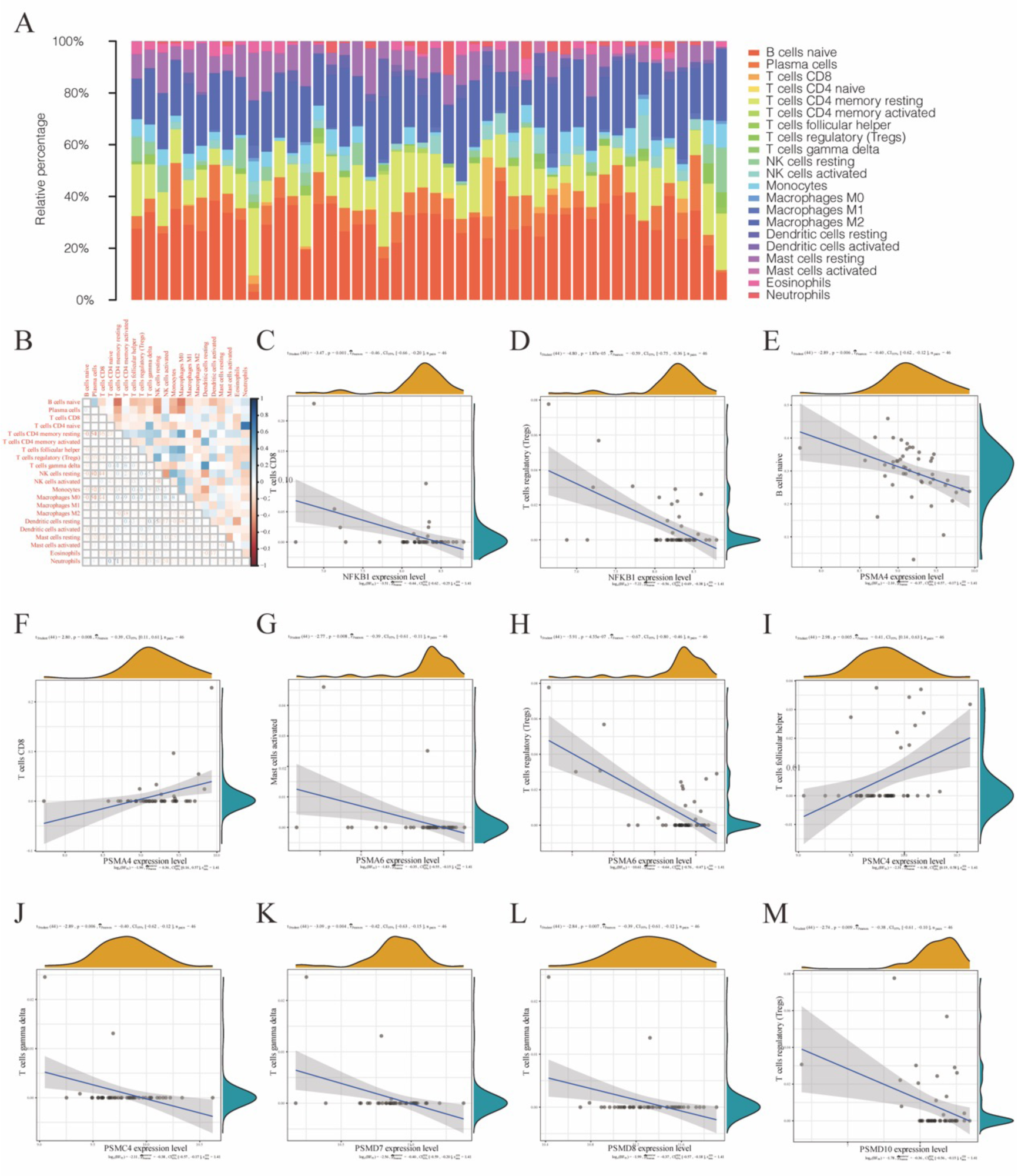
CIBERSORT immune cell infiltration analysis and correlation analysis between hubgene and immune cells. A. Barplot showing the proportions of 22 immune cells in a sepsis sample. The column in the figure is a sample. B. Correlation heat map of 22 immune cells infiltration. Blue indicated positive correlation, red indicated negative correlation, the darker the color, the stronger the correlation. C. The correlation between NFKB1 and CD8^+^ T cells. D. The correlation between NFKB1 and T regulatory cells. E. The correlation between PSMA4 and B naïve cells. F. The correlation between PSMA4 and CD8^+^ T cells. G. The correlation between PSMA6 and mast cells. H. The correlation between PSMA6 and T regulatory cells. I. The correlation between PSMC4 and T follicular helper cells. J. The correlation between PSMC4 and T gamma delta cells. K. The correlation between PSMD7 and T gamma delta cells. L. The correlation between PSMD8 and T gamma delta cells. M. The correlation between PSMD10 and T regulatory cells.

### 3.5 Hubgene subtype construction

When k = 2, the classification was reliable and stable (Figure 11A∼E). We divided the samples into GeneClusterA and GeneClusterB. And through principal component analysis, it was found that the classification was significantly different (Figure 11F). Among 10 hubgenes, NFKB1, PSMA4, and PSMA6 were dys-regulated between GeneClusterA and GeneClusterB (*P* < 0.001, < 0.01, and < 0.001, respectively) (Figure 11G).

**Figure 11.**
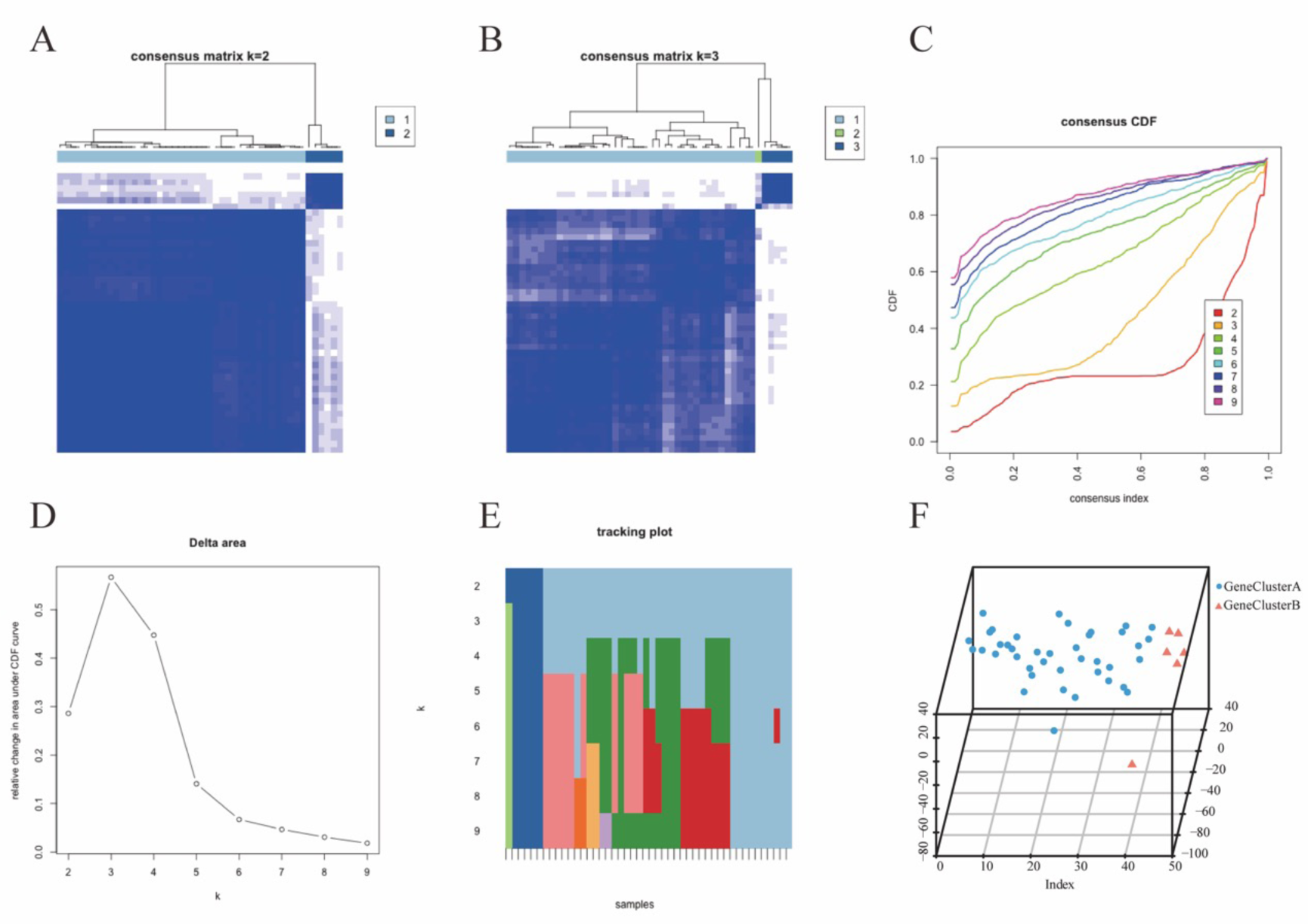
Hubgene subtype construction. A. Matrix heatmap for k = 2. B. Matrix heatmap for k = 3. C. Consistent cumulative distribution function plot. This figure allows a user to determine at what number of clusters, k, the cumulative distribution function reaches an approximate maximum, thus consensus and cluster con dence is at a maximum at this k. D. Delta area plot. The delta area score (y-axis) indicates the relative increase in cluster stability. E. Tracking plot. This plot provides a view of item cluster membership across different k and enables a user to track the history of clusters relative to earlier clusters. F. Principal component analysis. G. Difference analysis of top 10 hubgenes in hubgene subtype classification.

### 3.6 Experimental Validation

Relative mRNA levels of hypertrophy marker genes (ANP, BNP, and MYH7) and Hnrnpc, Fmr1, Fto in heart tissues from mice at four weeks after sham operations or TAC surgery (n=6). For statistical analysis, nonparametric statistical analysis was performed using the Mann-Whitney test for two groups.

## 4. Discussion

HCM is mainly caused by the mutations in sarcomere genes[45]. Relevant genetic testing has been recommended by guidelines and has promoted the diagnosis of HCM to a certain extent. However, more and more studies show that the clinical characteristics observed in patients with HCM is not all caused by abnormal sarcomeric proteins[2, 46, 47]. Therefore, it is necessary to explore alternative molecular mechanisms of HCM and identify more valuable biomarkers by utilizing contemporary methods to analyze biological complexity[48]. Here, we firstly identified 20 m6A-related regulators in HCM and constructed the m6A, immune, and hubgene subtypes. Finally, the key findings were validated the molecular biology experiments.

m6A is required for the formation of a nuclear body mediated by phase separation that maintains mRNA stability[49]. YTHDC1 mediates export of methylated mRNA from the nucleus to the cytoplasm and knockdown of YTHDC1 results in an extended residence time for nuclear m6A-containing mRNA, with an accumulation of transcripts in the nucleus and accompanying depletion within the cytoplasm[50]. YTHDC2 is a powerful endogenous ferroptosis inducer and targets SLC3A2 and SLC7A11[51, 52]. HNRNPC is attributed to the function of controlling the endogenous double-stranded RNA and the downstream interferon response[53]. Sequence-specific mRNAs instruct FMR1-ribonucleoprotein granules to undergo a dynamic phase switch, thus contributing to maternal mRNA decay. FTO mediated the m6A level of LINC00022 and MALAT to promote the tumorigenesis[54, 55]. WTAP-guided m6A modification contributes to the progression of hepatocellular carcinoma via the HuR-ETS1-p21/p27 axis[56]. IGF2BP2/3 m6A promotes vasculogenic mimicry in colorectal cancer via PI3K/AKT and ERK1/2 signaling[57] and attenuates the detrimental effect of irradiation on lung adenocarcinoma[58]. Here, we found that these m6A-related regulators were differentially expressed in HCM. Moreover, we illustrated the correlations of certain m6A-related regulators. Then we built the high-fidelity random forest model and used the top 5 important m6A-related regulators in HCM (namely FTO, FMR1, IGF2BP3, YTHDC1, and IGF2BP2) to construct the nomogram, which is a good predictor for the risk of HCM. According to the expression of these 5 m6A-related regulators, we constructed the m6A subtype. Based on the 329 differential genes of the two subtypes, we enriched many biological processes and signaling pathways known to be associated with HCM. Activation of CaMK-signaling pathway may be involved in the pathophysiology of HCM[59]. Post-translational activation of the CaMKII pathway is specific to sarcomere mutation-positive HCM, whereas sarcoplasmic endoplasmic reticular calcium ATPase 2 abundance and sarcoplasmic reticulum Ca^2+^ uptake are depressed in both sarcomere mutation-positive and -negative HCM[60]. HCM is characterized by enhanced oxidative stress, which achieves its highest values in the presence of LVOT obstruction in HCM patients[61] and reducing oxidative stress can be a viable therapeutic approach to attenuating the severity of cardiac dysfunction in HCM and prevent its progression[62]. Importantly, we also enriched oxidative stress-related signaling pathways. After protein-protein interaction of 329 differentially expressed genes, Hubgene-chemical network, Hubgene-microRNA network, and Hubgene-transcription factor network of the top 10 hubgenes among 329 differentially expressed genes were sequentially constructed, which not only provide ideas for the pathogenesis of HCM, but also give insights into the treatment of HCM.

m6A is closely connected to immune cell infiltration[20, 21] and the immune system engage in the HCM mechanisms[22, 23]. After m6A subtype, we constructed immune subtype according to the immune cell infiltration levels. We demonstrated the correlation of some differential genes with immune cell infiltration and various immune cells, which further deepens the understanding of immunity in the occurrence and development of HCM. And then hubgene subtype classification was constructed based on the expression of top 10 hubgenes and NFKB1, PSMA4, and PSMA6 were dys-regulated between hubgene subtypes.

This study establishes a clinically accessible model for predicting HCM risk through bioinformatics analysis using transcriptome datasets of HCM patients and healthy controls, and furthers the understanding of the molecular mechanisms of m6A and immunity in HCM. Nonetheless, our study still has some limitations and inadequacies. Firstly, although we used different datasets for the analysis, because this was a reanalysis based on the existing data, we could not fully obtain more pathological, clinical and prognostic information of the dataset, which led us to build a risk model later, rather than provide more evidence for treatment or prognosis. In addition, we constructed different networks, which have a certain role in promoting the pathogenesis and clinical treatment of HCM. Although we have carried out biological experiments to verify important results, most of the results we obtained are based on computer analysis. Many basic experiments and clinical experiments are warranted to validate our findings.

## 5. Conclusion

The systematic analysis of the transcriptional profiles of HCM and exploration of the potential underlying molecular mechanisms led to the identification of m6A, immune and hubgene subtype of HCM. Based on these, a risk assessment model of HCM was developed that brings a novel understanding to the current understanding of HCM.

## Abbreviations

## Declarations

### Ethics approval and consent to participate

All animal experiment was conducted in accordance with the internationally accepted principles for laboratory animal use and care and was reviewed and approved by the Experimental Animal Center of Zhengzhou University (Zhengzhou, China) (ZZU-LAC20220311[23]). Our study is reported in accordance with the Animal Research: Reporting of *in vivo* Experiments guidelines.

### Consent for publication

Not applicable.

### Availability of data and materials

All data generated or analyzed during this study are included in this published article

and its supplementary information files.

### Authors contributions

BL and NG conceived, designed, and planned the study. BL and NG acquired and analyzed the data. BL completed the experiments. BL and NG interpreted the results. BL drafted the manuscript and NG contributed to the critical revision of the manuscript. All authors read and approved the final manuscript.

## Data Availability

All data produced in the present work are contained in the manuscript

## Acknowledgments

We are grateful to all research scientists who participated in the aforementioned databases.

---

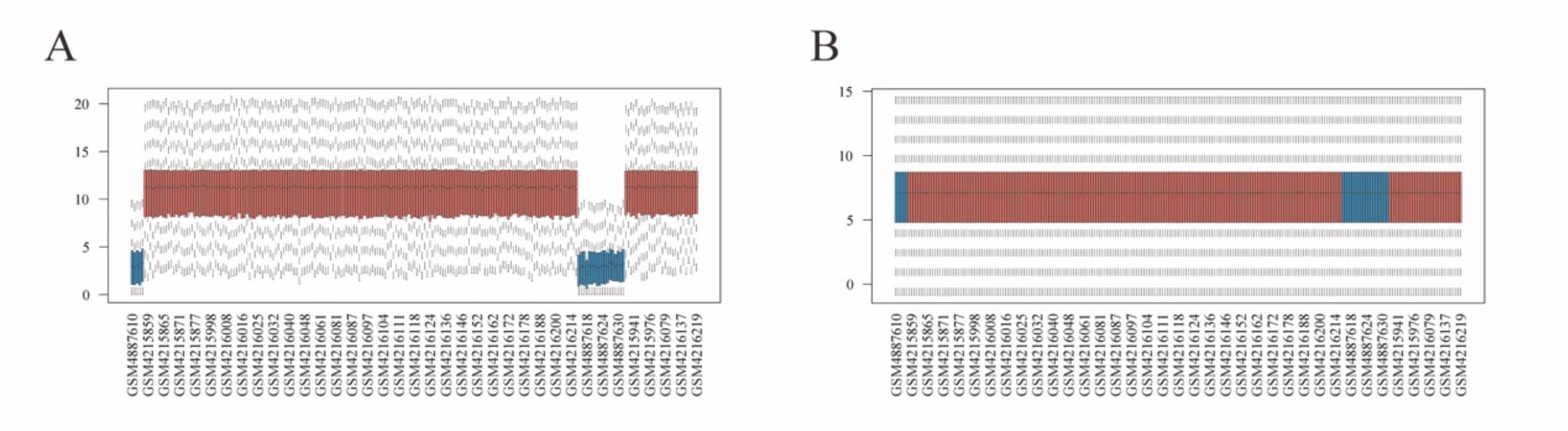

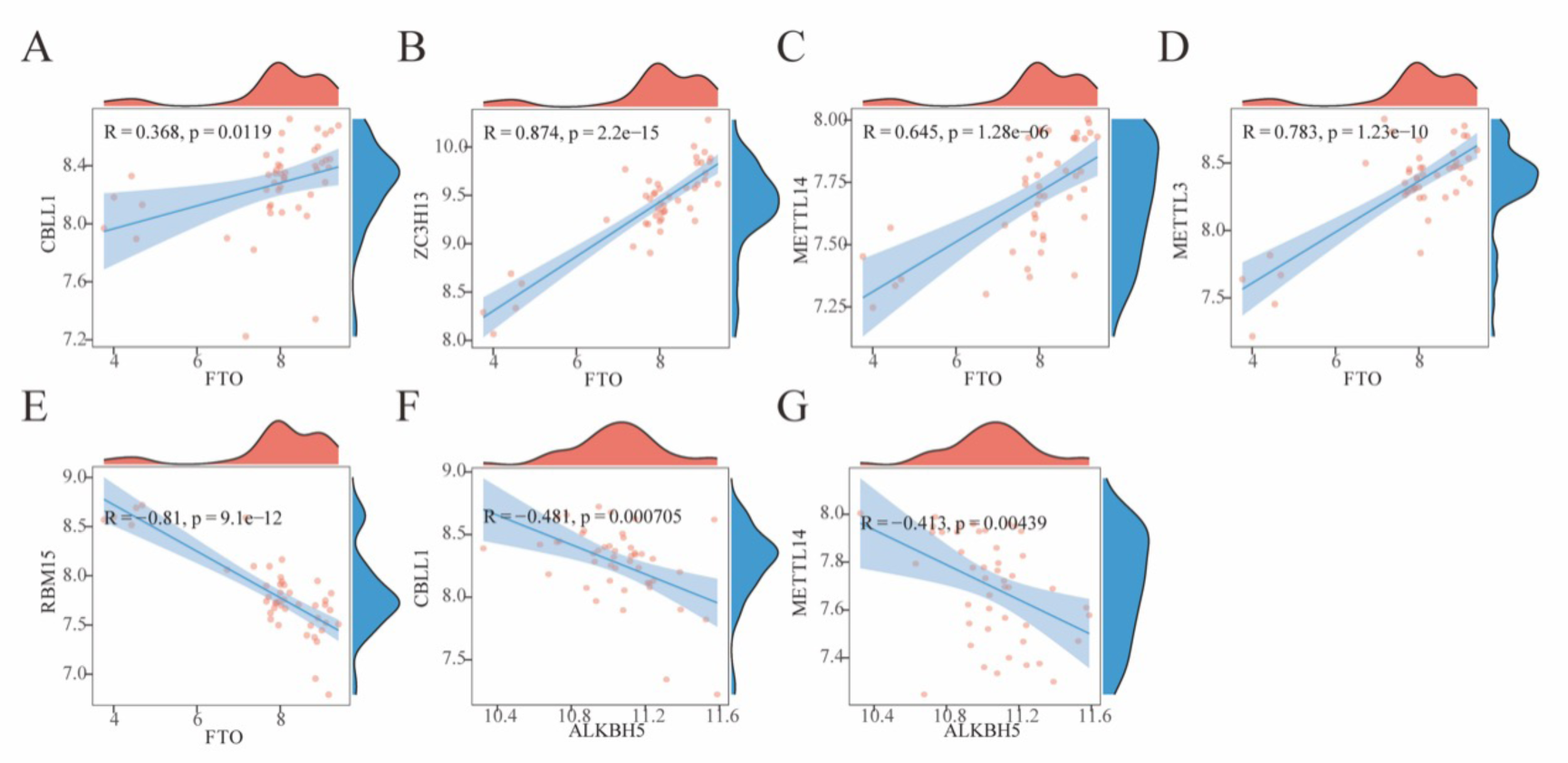

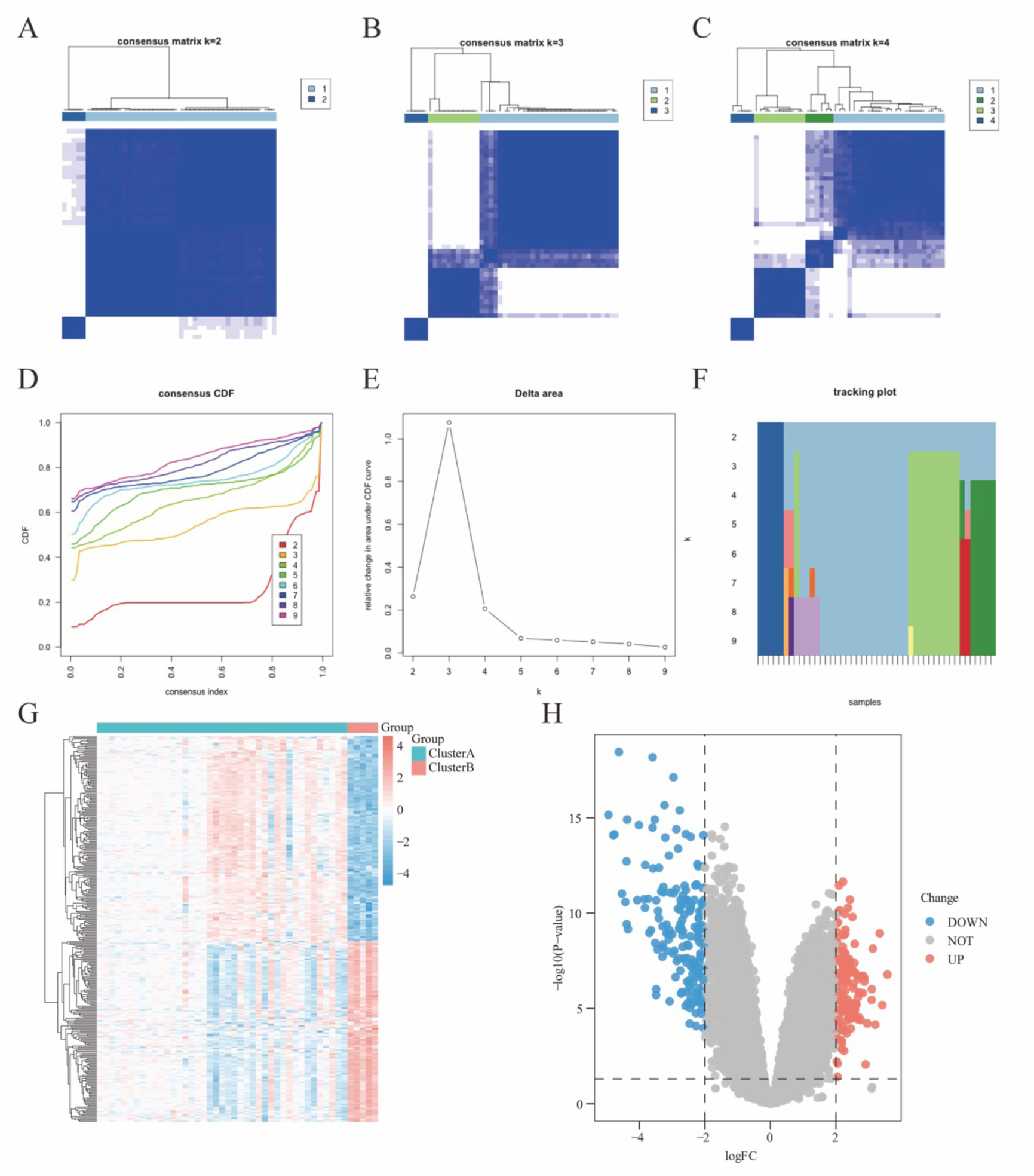

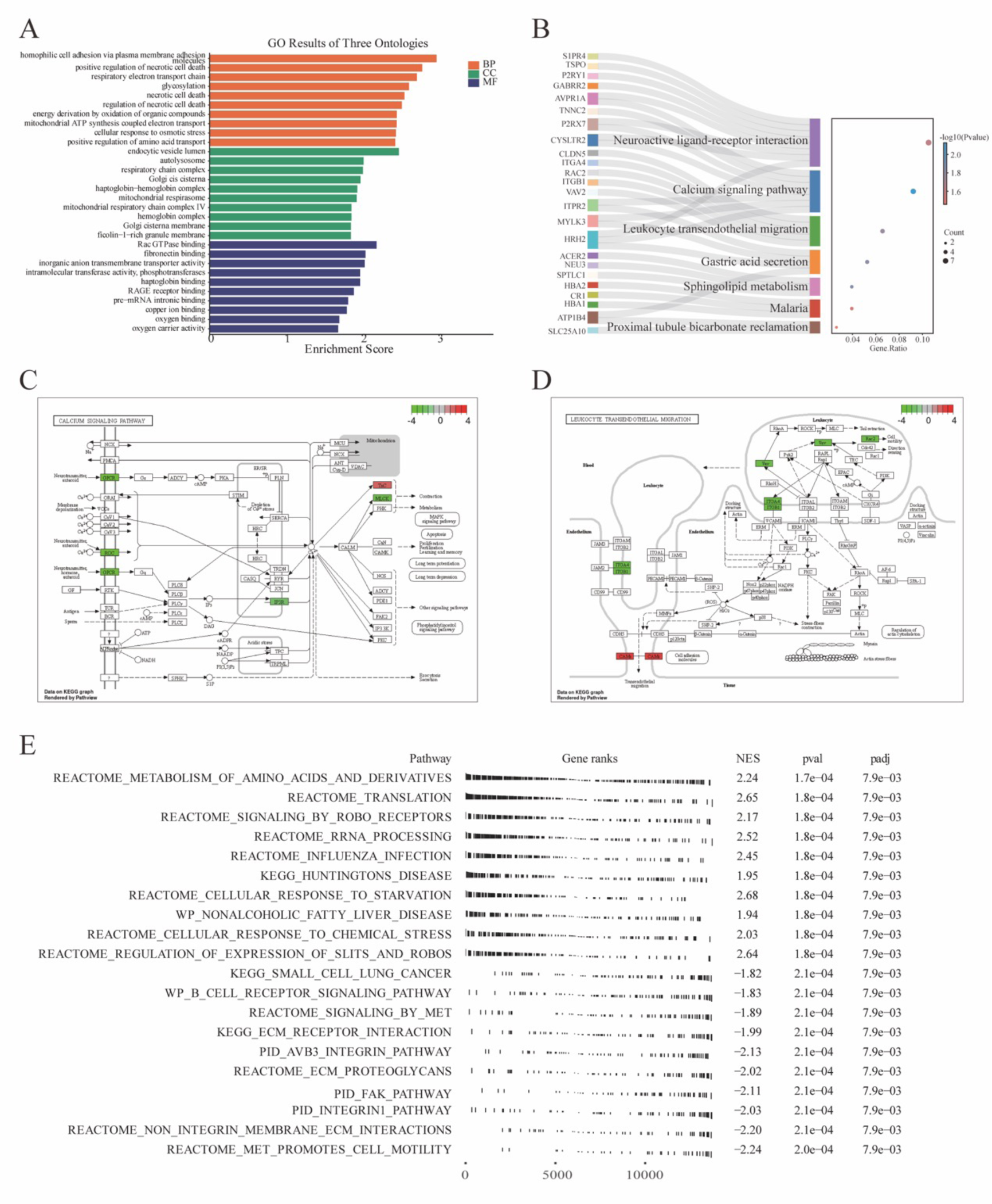

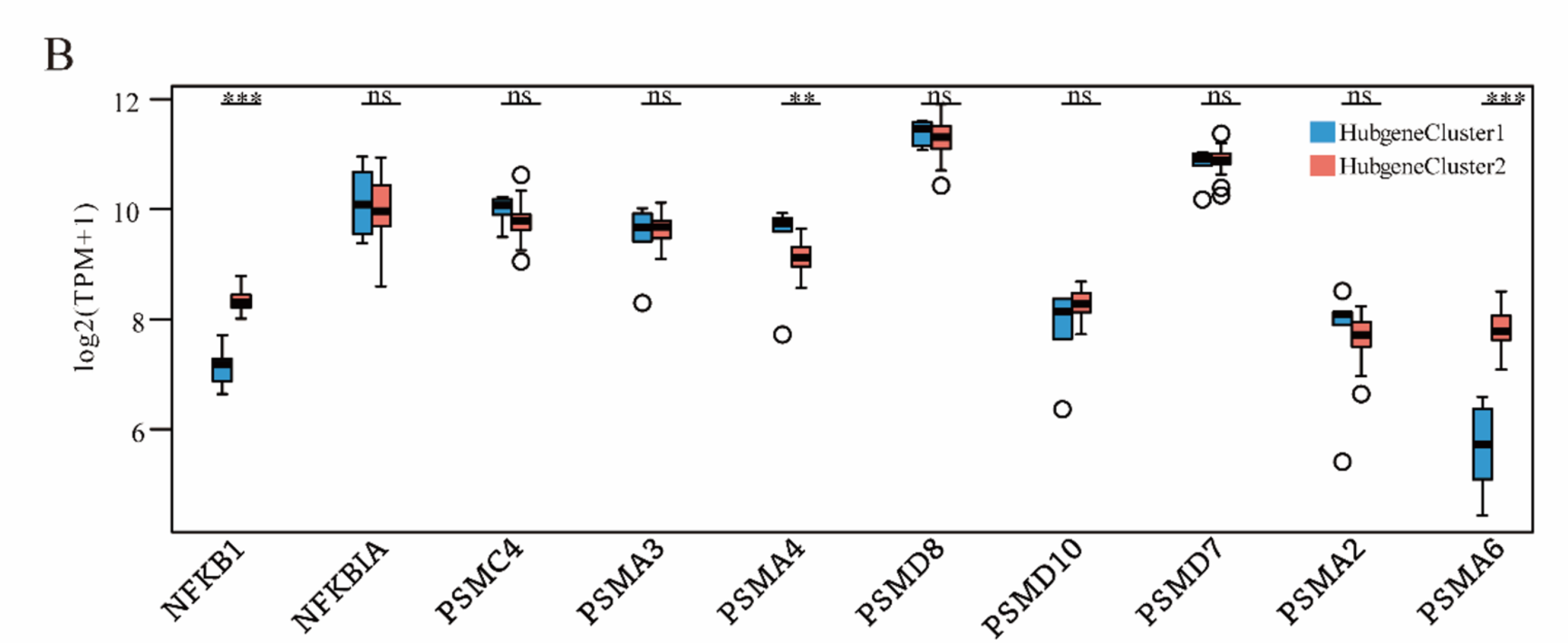

## Additional files

Table 1. Details of datasets.

**Table S1.** Primer sequences.

**Figure S1.**
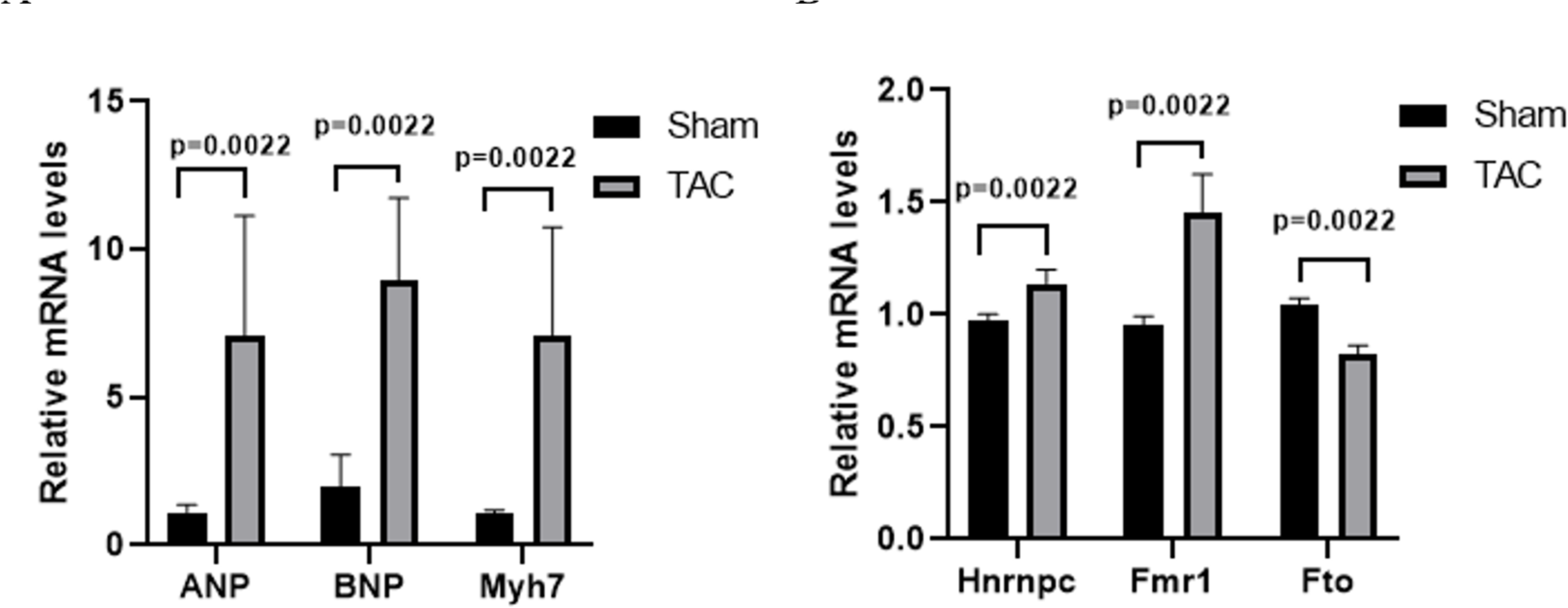
Datasets collation. A. All samples were pooled before removing batch effects. B. All samples were pooled after removing batch effects.

## References

1. Palandri C, Santini L, Argirò A, Margara F, Doste R, Bueno-Orovio A, Olivotto I, Coppini R: Pharmacological Management of Hypertrophic Cardiomyopathy: From Bench to Bedside. Drugs 2022, 82(8):889–912.

2. Ommen SR, Mital S, Burke MA, Day SM, Deswal A, Elliott P, Evanovich LL, Hung J, Joglar JA, Kantor P et al: 2020 AHA/ACC Guideline for the Diagnosis and Treatment of Patients With Hypertrophic Cardiomyopathy: A Report of the American College of Cardiology/American Heart Association Joint Committee on Clinical Practice Guidelines. Circulation 2020, 142(25):e558–e631.

3. Elliott PM, Anastasakis A, Borger MA, Borggrefe M, Cecchi F, Charron P, Hagege AA, Lafont A, Limongelli G, Mahrholdt H et al: 2014 ESC Guidelines on diagnosis and management of hypertrophic cardiomyopathy: the Task Force for the Diagnosis and Management of Hypertrophic Cardiomyopathy of the European Society of Cardiology (ESC). European heart journal 2014, 35(39):2733–2779.

4. Covella M, Rowin EJ, Hill NS, Preston IR, Milan A, Opotowsky AR, Maron BJ, Maron MS, Maron BA: Mechanism of Progressive Heart Failure and Significance of Pulmonary Hypertension in Obstructive Hypertrophic Cardiomyopathy. Circ Heart Fail 2017, 10(4):e003689.

5. Ommen SR, Semsarian C: Hypertrophic cardiomyopathy: a practical approach to guideline directed management. Lancet (London, England) 2021, 398(10316):2102–2108.

6. Geisterfer-Lowrance AA, Christe M, Conner DA, Ingwall JS, Schoen FJ, Seidman CE, Seidman JG: A mouse model of familial hypertrophic cardiomyopathy. Science (New York, NY) 1996, 272(5262):731-734.

7. Pagiatakis C, Di Mauro V: The Emerging Role of Epigenetics in Therapeutic Targeting of Cardiomyopathies. Int J Mol Sci 2021, 22(16):8721.

8. Takeda Y, Demura M, Yoneda T, Takeda Y: DNA Methylation of the Angiotensinogen Gene, AGT, and the Aldosterone Synthase Gene, CYP11B2 in Cardiovascular Diseases. Int J Mol Sci 2021, 22(9):4587.

9. Liu C, Gu L, Deng W-J, Meng Q-C, Li N, Dai G, Yu S, Fang H: N6-Methyladenosine RNA Methylation in Cardiovascular Diseases. Frontiers in cardiovascular medicine 2022, 9:887838.

10. Chien C-S, Li JY-S, Chien Y, Wang M-L, Yarmishyn AA, Tsai P-H, Juan C- C, Nguyen P, Cheng H-M, Huo T-I et al: METTL3-dependent N(6)-methyladenosine RNA modification mediates the atherogenic inflammatory cascades in vascular endothelium. Proc Natl Acad Sci U S A 2021, 118(7):e2025070118.

11. Li B-C, Zhang T, Liu M-X, Cui Z, Zhang Y-H, Liu M, Liu Y, Sun Y, Li M, Tian Y et al: RNA N(6)-methyladenosine modulates endothelial atherogenic responses to disturbed flow in mice. eLife 2022, 11:e69906.

12. Su X, Shen Y, Jin Y, Kim I-M, Weintraub NL, Tang Y: Aging-Associated Differences in Epitranscriptomic m6A Regulation in Response to Acute Cardiac Ischemia/Reperfusion Injury in Female Mice. Front Pharmacol 2021, 12:654316.

13. Mo X-B, Lei S-F, Zhang Y-H, Zhang H: Examination of the associations between m(6)A-associated single-nucleotide polymorphisms and blood pressure. Hypertension research : official journal of the Japanese Society of Hypertension 2019, 42(10):1582–1589.

14. Cheng P-K, Han H-K, Chen F-L, Cheng L, Ma C, Huang H, Chen C, Li H, Cai H, Huang H et al: Amelioration of acute myocardial infarction injury through targeted ferritin nanocages loaded with an ALKBH5 inhibitor. Acta biomaterialia 2022, 140:481–491.

15. Qian B-H, Wang P, Zhang D-H, Wu L: m6A modification promotes miR-133a repression during cardiac development and hypertrophy via IGF2BP2. Cell Death Discov 2021, 7:157–157.

16. Mathiyalagan P, Adamiak M, Mayourian J, Sassi Y, Liang Y, Agarwal N, Jha D, Zhang S, Kohlbrenner E, Chepurko E et al: FTO-Dependent N(6)-Methyladenosine Regulates Cardiac Function During Remodeling and Repair. Circulation 2019, 139(4):518–532.

17. Zhang B-J, Xu Y-M, Cui X-T, Jiang H, Luo W, Weng X, Wang Y, Zhao Y, Sun A, Ge J: Alteration of m6A RNA Methylation in Heart Failure With Preserved Ejection Fraction. Frontiers in cardiovascular medicine 2021, 8:647806.

18. Shulman Z, Stern-Ginossar N: The RNA modification N(6)-methyladenosine as a novel regulator of the immune system. Nature immunology 2020, 21(5):501–512.

19. Wang Y-N, Yu C-Y, Jin H-Z: RNA N(6)-Methyladenosine Modifications and the Immune Response. Journal of immunology research 2020, 2020:6327614.

20. Liang C-Z, Wang S, Zhang M, Li T: Diagnosis, clustering, and immune cell infiltration analysis of m6A-related genes in patients with acute myocardial infarction-a bioinformatics analysis. Journal of thoracic disease 2022, 14(5):1607–1619.

21. Yin F-X, Zhang H, Guo P-P, Wu Y, Zhao X, Li F, Bian C, Chen C, Han Y, Liu K: Comprehensive Analysis of Key m6A Modification Related Genes and Immune Infiltrates in Human Aortic Dissection. Frontiers in cardiovascular medicine 2022, 9:831561.

22. Kardaszewicz B, Rogala E, Tendera M, Kardaszewicz P, Jarzab J: Circulating immune complexes in hypertrophic cardiomyopathy and ischemic heart disease. Kardiologia polska 1991, 34(1):21–24.

23. Lüscher TF: An update on cardiomyopathies: immune-mediated diseases, sarcoidosis, and peripartum and hypertrophic cardiomyopathies during pregnancy. European heart journal 2017, 38(35):2635–2638.

24. Wang Z-Q, Ruan H-Y, Li L-Y, Wei X, Zhu Y, Wei J, Chen X, He S: Assessing the relationship between systemic immune-inflammation index and mortality in patients with hypertrophic cardiomyopathy. Upsala journal of medical sciences 2021, 126.

25. Maron BA, Wang R-S, Shevtsov S, Drakos SG, Arons E, Wever-Pinzon O, Huggins GS, Samokhin AO, Oldham WM, Aguib Y et al: Individualized interactomes for network-based precision medicine in hypertrophic cardiomyopathy with implications for other clinical pathophenotypes. Nat Commun 2021, 12(1):873–873.

26. Zhou J-G, Liang B, Jin S-H, Liao H-L, Du G-B, Cheng L, Ma H, Gaipl US: Development and Validation of an RNA-Seq-Based Prognostic Signature in Neuroblastoma. Front Oncol 2019, 9:1361.

27. Zhou J-G, Liang B, Liu J-G, Jin S-H, He S-S, Frey B, Gu N, Fietkau R, Hecht M, Ma H et al: Identification of 15 lncRNAs Signature for Predicting Survival Benefit of Advanced Melanoma Patients Treated with Anti-PD-1 Monotherapy. Cells 2021, 10(5):977.

28. Kainuma A, Ning Y, Kurlansky PA, Wang AS, Latif F, Sayer GT, Uriel N, Kaku Y, Naka Y, Takeda K: Predictors of one-year outcome after cardiac re-transplantation: Machine learning analysis. Clinical transplantation 2022:e14761.

29. Lu W-C, Chen H, Liang B, Ou C-P, Zhang M, Yue Q, Xie J: Integrative Analyses and Verification of the Expression and Prognostic Significance for RCN1 in Glioblastoma Multiforme. Front Mol Biosci 2021, 8:736947.

30. Zhang M-W, Chen H, Liang B, Wang X-Z, Gu N, Xue F-Q, Yue Q-Y, Zhang Q-Y, Hong J-S: Prognostic Value of mRNAsi/Corrected mRNAsi Calculated by the One-Class Logistic Regression Machine-Learning Algorithm in Glioblastoma Within Multiple Datasets. Front Mol Biosci 2021, 8:777921.

31. Wilkerson MD, Hayes DN: ConsensusClusterPlus: a class discovery tool with confidence assessments and item tracking. Bioinformatics 2010, 26(12):1572–1573.

32. Yu G-C, Wang L-G, Han Y, He Q-Y: clusterProfiler: an R package for comparing biological themes among gene clusters. Omics : a journal of integrative biology 2012, 16(5):284–287.

33. Luo W-J, Brouwer C: Pathview: an R/Bioconductor package for pathway-based data integration and visualization. Bioinformatics 2013, 29(14):1830–1831.

34. Liberzon A, Birger C, Thorvaldsdóttir H, Ghandi M, Mesirov JP, Tamayo P: The Molecular Signatures Database (MSigDB) hallmark gene set collection. Cell Syst 2015, 1(6):417–425.

35. Subramanian A, Tamayo P, Mootha VK, Mukherjee S, Ebert BL, Gillette MA, Paulovich A, Pomeroy SL, Golub TR, Lander ES et al: Gene set enrichment analysis: a knowledge-based approach for interpreting genome-wide expression profiles. Proc Natl Acad Sci U S A 2005, 102(43):15545–15550.

36. Szklarczyk D, Gable AL, Nastou KC, Lyon D, Kirsch R, Pyysalo S, Doncheva NT, Legeay M, Fang T, Bork P et al: The STRING database in 2021: customizable protein-protein networks, and functional characterization of user-uploaded gene/measurement sets. Nucleic acids research 2021, 49(D1):D605–D612.

37. Chin C-H, Chen S-H, Wu H-H, Ho C-W, Ko M-T, Lin C-Y: cytoHubba: identifying hub objects and sub-networks from complex interactome. BMC systems biology 2014, 8 Suppl 4(Suppl 4):S11.

38. Shannon P, Markiel A, Ozier O, Baliga NS, Wang JT, Ramage D, Amin N, Schwikowski B, Ideker T: Cytoscape: a software environment for integrated models of biomolecular interaction networks. Genome Res 2003, 13(11):2498–2504.

39. Zhou G-Y, Soufan O, Ewald J, Hancock REW, Basu N, Xia J-G: NetworkAnalyst 3.0: a visual analytics platform for comprehensive gene expression profiling and meta-analysis. Nucleic acids research 2019, 47(W1):W234–w241.

40. Huang H-Y, Lin Y-C-D, Li J, Huang K-Y, Shrestha S, Hong H-C, Tang Y, Chen Y-G, Jin C-N, Yu Y et al: miRTarBase 2020: updates to the experimentally validated microRNA-target interaction database. Nucleic acids research 2020, 48(D1):D148–D154.

41. Newman AM, Liu CL, Green MR, Gentles AJ, Feng W, Xu Y, Hoang CD, Diehn M, Alizadeh AA: Robust enumeration of cell subsets from tissue expression profiles. Nat Methods 2015, 12(5):453–457.

42. Rockman HA, Ross RS, Harris AN, Knowlton KU, Steinhelper ME, Field LJ, Ross J, Jr., Chien KR: Segregation of atrial-specific and inducible expression of an atrial natriuretic factor transgene in an in vivo murine model of cardiac hypertrophy. Proc Natl Acad Sci U S A 1991, 88(18):8277–8281.

43. Liang B, Zhang X-X, Li R, Gu N: Guanxin V protects against ventricular remodeling after acute myocardial infarction through the interaction of TGF-β1 and Vimentin. Phytomedicine 2022, 95:153866.

44. Liang B, Zhang X-X, Li R, Zhu Y-C, Tian X-J, Gu N: Guanxin V alleviates acute myocardial infarction by restraining oxidative stress damage, apoptosis, and fibrosis through the TGF-β1 signalling pathway. Phytomedicine 2022, 100:154077.

45. van der Velde N, Huurman R, Hassing HC, Budde RPJ, van Slegtenhorst MA, Verhagen JMA, Schinkel AFL, Michels M, Hirsch A: Novel Morphological Features on CMR for the Prediction of Pathogenic Sarcomere Gene Variants in Subjects Without Hypertrophic Cardiomyopathy. Frontiers in cardiovascular medicine 2021, 8:727405.

46. Bos JM, Will ML, Gersh BJ, Kruisselbrink TM, Ommen SR, Ackerman MJ: Characterization of a phenotype-based genetic test prediction score for unrelated patients with hypertrophic cardiomyopathy. Mayo Clinic proceedings 2014, 89(6):727–737.

47. Neubauer S, Kolm P, Ho CY, Kwong RY, Desai MY, Dolman SF, Appelbaum E, Desvigne-Nickens P, DiMarco JP, Friedrich MG et al: Distinct Subgroups in Hypertrophic Cardiomyopathy in the NHLBI HCM Registry. Journal of the American College of Cardiology 2019, 74(19):2333–2345.

48. Zheng X-F, Liu G-Y, Huang R-N: Identification and Verification of Feature Immune-Related Genes in Patients With Hypertrophic Cardiomyopathy Based on Bioinformatics Analyses. Frontiers in cardiovascular medicine 2021, 8:752559.

49. Cheng Y-M, Xie W, Pickering BF, Chu KL, Savino AM, Yang X, Luo H, Nguyen DT, Mo S, Barin E et al: N(6)-Methyladenosine on mRNA facilitates a phase-separated nuclear body that suppresses myeloid leukemic differentiation. Cancer cell 2021, 39(7):958–972.e958.

50. Roundtree IA, Luo G-Z, Zhang Z-J, Wang X, Zhou T, Cui Y, Sha J, Huang X, Guerrero L, Xie P et al: YTHDC1 mediates nuclear export of N(6)-methyladenosine methylated mRNAs. eLife 2017, 6:e31311.

51. Ma L-F, Chen T-X, Zhang X, Miao Y, Tian X, Yu K, Xu X, Niu Y, Guo S, Zhang C et al: The m(6)A reader YTHDC2 inhibits lung adenocarcinoma tumorigenesis by suppressing SLC7A11-dependent antioxidant function. Redox biology 2021, 38:101801.

52. Ma L-F, Zhang X, Yu K-K, Xu X, Chen T, Shi Y, Wang Y, Qiu S, Guo S, Cui J et al: Targeting SLC3A2 subunit of system X(C)(-) is essential for m(6)A reader YTHDC2 to be an endogenous ferroptosis inducer in lung adenocarcinoma. Free radical biology & medicine 2021, 168:25–43.

53. Wu Y-S, Zhao W-W, Liu Y, Tan X, Li X, Zou Q, Xiao Z, Xu H, Wang Y, Yang X: Function of HNRNPC in breast cancer cells by controlling the dsRNA-induced interferon response. The EMBO journal 2018, 37(23):e99017.

54. Cui Y-B, Zhang C-Y, Ma S-S, Li Z, Wang W, Li Y, Ma Y, Fang J, Wang Y, Cao W et al: RNA m6A demethylase FTO-mediated epigenetic up-regulation of LINC00022 promotes tumorigenesis in esophageal squamous cell carcinoma. Journal of experimental & clinical cancer research : CR 2021, 40(1):294.

55. Tao L, Mu X-Y, Chen H-G, Jin D, Zhang R, Zhao Y, Fan J, Cao M, Zhou Z: FTO modifies the m6A level of MALAT and promotes bladder cancer progression. Clinical and translational medicine 2021, 11(2):e310.

56. Chen Y-H, Peng C-H, Chen J-R, Chen D, Yang B, He B, Hu W, Zhang Y, Liu H, Dai L et al: WTAP facilitates progression of hepatocellular carcinoma via m6A-HuR-dependent epigenetic silencing of ETS1. Molecular cancer 2019, 18(1):127.

57. Liu X, He H-J, Zhang F-W, Hu X, Bi F, Li K, Yu H, Zhao Y, Teng X, Li J et al: m6A methylated EphA2 and VEGFA through IGF2BP2/3 regulation promotes vasculogenic mimicry in colorectal cancer via PI3K/AKT and ERK1/2 signaling. Cell death & disease 2022, 13(5):483.

58. Hao C-C, Xu C-Y, Zhao X-Y, Luo J-N, Wang G, Zhao L-H, Ge X, Ge X-F: Up-regulation of VANGL1 by IGF2BPs and miR-29b-3p attenuates the detrimental effect of irradiation on lung adenocarcinoma. Journal of experimental & clinical cancer research : CR 2020, 39(1):256.

59. Chen P, Li Z, Nie J, Wang H, Yu B, Wen Z, Sun Y, Shi X, Jin L, Wang D-W: MYH7B variants cause hypertrophic cardiomyopathy by activating the CaMK-signaling pathway. Science China Life sciences 2020, 63(9):1347–1362.

60. Helms AS, Alvarado FJ, Yob J, Tang VT, Pagani F, Russell MW, Valdivia HH, Day SM: Genotype-Dependent and -Independent Calcium Signaling Dysregulation in Human Hypertrophic Cardiomyopathy. Circulation 2016, 134(22):1738–1748.

61. Dimitrow PP, Undas A, Wołkow P, Tracz W, Dubiel JS: Enhanced oxidative stress in hypertrophic cardiomyopathy. Pharmacological reports : PR 2009, 61(3):491–495.

62. Hassoun R, Budde H, Zhazykbayeva S, Herwig M, Sieme M, Delalat S, Mostafi N, Gömöri K, Tangos M, Jarkas M et al: Stress activated signalling impaired protein quality control pathways in human hypertrophic cardiomyopathy. International journal of cardiology 2021, 344:160–169.

